# Using Explainable AI to Identify Disease-Relevant and Deep Brain Stimulation Treatment-Sensitive Gait Features in Parkinson’s Disease

**DOI:** 10.1101/2025.11.27.25341152

**Authors:** Zhongke Mei, Alain Ryser, Gianluca Amprimo, Jinhao Wang, Julia Vogt, Deepak K. Ravi

**Author notes:** This work involved human subjects or animals in its research. Approval of all ethical and experimental procedures and protocols was granted by the Zurich Cantonal Ethics Commission and carried out in accordance with the Declaration of Helsinki, the guidelines of Good Clinical Practice, and the Swiss regulatory authority’s requirements. Application No. 201500141.

## Abstract

Gait impairment is a characteristic motor deficit of Parkinson’s disease (PD) and a critical but insufficiently understood target of deep brain stimulation (DBS). Identifying robust gait biomarkers that capture both disease-related deficits and stimulation-induced improvements remains a major challenge. In this study, we analyzed 35 spatiotemporal gait parameters from individuals with PD before and after subthalamic DBS, alongside age-matched healthy controls, during continuous walking. Multiple machine learning classifiers were evaluated, with XGBoost achieving the best discrimination between groups. To ensure interpretability, we employed grouped SHapley Additive exPlanations (SHAP), which ranked feature importance while mitigating redundancy from correlated parameters. Feature selection consistently emphasized step width variability, step width asymmetry, bilateral interlimb coordination, and the anteroposterior margin of stability. Importantly, a compact set of five overlapping features after selection not only distinguished PD from healthy gait but also shifted toward healthy ranges after DBS. Unlike conventional mean-based metrics, the selected characteristics provided robust markers of both pathology and treatment response. Our findings demonstrate that explainable AI can identify physiologically grounded gait features that serve as biomarkers for both PD severity and DBS responsiveness, supporting more precise evaluation of neuromodulation outcomes and individualized patient management.

## I. INTRODUCTION

Parkinson’s disease (PD) is a progressive neurodegenerative disorder primarily characterized by the degeneration of dopaminergic neurons within the substantia nigra pars compacta [1]. PD is notably the most common age-related motoric neurodegenerative disease, with prevalence rates reported to range from 108 to 257 per 100,000 individuals in Europe [1, 2]. The motor symptoms of PD are diverse, including tremor, rigidity, bradykinesia, and postural instability [3–5]. Gait disturbances and postural instability are particularly significant challenges in PD, affecting mobility, independence, and quality of life [6–8]. Gait impairments in PD commonly include reduced gait speed and step length, as well as impaired rhythmicity of walking patterns [9, 10], frequently accompanied by asymmetric lower limb movements [11, 12] and reduced bilateral coordination [13, 14]. Levodopa is the gold standard for managing motor symptoms in PD [15]. However, its effects are limited to motor improvement only [16]. Moreover, the effectiveness of the medication may diminish over time with the disease progression, which leads to motor fluctuations and the onset of medication-related complications such as dyskinesia [17–19].

Deep brain stimulation (DBS) has become an essential therapeutic option for addressing motor symptoms in PD, especially for patients experiencing limited benefits from medication, such as those with persistent tremors [20, 21]. This surgical technique involves precisely placing electrodes into targeted regions of the brain, with the subthalamic nucleus (STN-DBS) being a primary target due to its critical role in regulating motor functions [22]. These electrodes deliver high-frequency electrical stimulation, modulating abnormal neural patterns and restoring functional balance in motor circuits [23]. Consequently, STN-DBS provides sustained symptom relief, diminishes motor fluctuations, and significantly enhances patients’ quality of life [24–26].

Even though STN-DBS is well established as an effective treatment for managing motor symptoms in PD, its benefits have been primarily demonstrated through reductions in scores on Part III of the Unified Parkinson’s Disease Rating Scale (UPDRS), which evaluates core motor signs such as tremor, rigidity, and bradykinesia [27–30]. However, gait, a key determinant of functional mobility and independence, is only minimally represented in this clinical assessment. As a result, the specific effects of STN-DBS on gait remain less well understood. This limited representation may lead to an underestimation of gait-related impairments during clinical evaluation and surgical planning, potentially contributing to the variability in treatment outcomes observed among people with PD [31–33].

Recent advancements in gait analysis, including optical motion capture systems and wearable sensors, have allowed quantification of detailed spatiotemporal characteristics of gait [34–36]. Quantitative assessments have highlighted specific alterations due to the PD in gait parameters, such as increased gait variability, increased cadence, reduced stride length, reduced swing duration, and prolonged stance duration, even in early disease stages [8, 36–38]. The quantification of spatiotemporal gait parameters is also employed in evaluating the effectiveness of DBS treatments in PD. Several studies have demonstrated the beneficial effects of STN-DBS on gait parameters, such as increased stride length, improved gait velocity, reduced double-stance duration, and decreased gait variability [39–44]. Conversely, other investigations reported that STN-DBS could exacerbate certain gait impairments, including increased gait asymmetry, reduced gait speed, shorter step lengths, increased cadence, and heightened gait dyscoordination [27, 45–47].

Despite increasing interest in utilizing gait metrics to evaluate PD, their interpretation remains challenging due to the complex and multifactorial nature of gait. Many spatiotemporal gait features are influenced not only by disease severity but also by stimulation settings, compensatory mechanisms, and other individual-specific factors [46, 48, 49]. As a result, analyses that focus on isolated gait metrics may lead to oversimplified interpretations that fail to capture the complex interplay between underlying pathology and neuromodulation-induced changes [50]. To overcome this, there is a critical need to identify a concise and reliable subset of gait biomarkers, defined as parameters that exhibit consistent, reproducible associations with clinical severity or therapeutic response across individuals and measurement contexts. However, this is complicated by redundancy among features, susceptibility to confounding, and the increased noise and computational complexity that arise from high-dimensional datasets. These challenges hinder the extraction of clinically meaningful insights and highlight the importance of reducing redundancy.

In recent years, machine learning (ML) techniques have emerged as powerful tools for navigating the complexity of high-dimensional gait datasets. These data-driven approaches not only enable the objective identification of key gait parameters that best discriminate between Parkinson’s disease (PD) and healthy gait patterns using algorithms such as Random Forest, Support Vector Machine, and Artificial Neural Networks [49, 51–57]. However, with the combination of explainable AI, the data-driven approach will also allow for transparent interpretation of model decisions. Explainable AI frameworks such as SHapley Additive exPlanations (SHAP) provide both global and individual-level quantification of feature contributions, making it possible to link algorithmic outputs to clinically meaningful gait characteristics. This transparency is critical for clinical adoption, as it enables healthcare professionals to understand why a model prioritizes certain parameters, thereby enhancing trust, interpretability, and the potential for integration into decision-support systems. Previous studies applying such approaches have repeatedly identified features such as step length, step width variability, stride length variability, double-support time variability, stance phase duration, and gait asymmetry as discriminative biomarkers for PD, underscoring the value of interpretable methods in highlighting clinically relevant gait metrics [49, 52, 56, 58].

Nevertheless, most existing studies have either focused on differentiating PD gait patterns from those of healthy controls or on predicting surgical outcomes using preoperative assessments alone [59]. As a result, there remains a critical lack of research aimed at systematically identifying gait parameters that are both characteristics of the motor impairments seen in individuals eligible for DBS and sensitive to DBS-induced changes. This gap limits our ability to establish gait biomarkers that are not only reflective of advanced disease features but also responsive to therapeutic treatment. Addressing this limitation is essential for advancing the development of reliable, objective markers of treatment efficacy and supporting more individualized DBS strategies in clinical practice.

In this study, we address this gap by leveraging quantitative gait data from individuals with PD who underwent STN-DBS. By analyzing gait patterns both before and after surgery and comparing them with data from age-matched healthy controls, we aimed to identify a concise set of gait parameters that reliably capture both disease-related impairments and treatment-induced changes. Using an explainable AI framework combining machine learning–based classification with SHAP-driven interpretability, we derived candidate gait biomarkers that may support objective evaluation of DBS effectiveness and inform more personalized management strategies in PD.

## II. Materials & Methods

### A. Data sets description

For this study, we used a previously collected dataset introduced by Mei et al. 2023 [60] and Amprimo et al. 2024 [59]. The dataset comprises 49 individuals diagnosed with Parkinson’s disease (PD), whose gait kinematics were recorded in their best medical state: ON-medication condition prior to STN-DBS electrode implantation (PRE), and in the combined ON-medication and ON-stimulation condition approximately six months post-surgery (POST) [60]. Ethical approval for data collection was granted by the Cantonal Ethics Commission Zurich (Protocol Number: 201500141), and all participants provided written informed consent. As detailed in prior studies using this dataset [27, 60], participants walked barefoot for 10 continuous minutes at a self-selected pace without any form of assistance. The walking task followed an “8”-shaped path marked by two signs placed 10 meters apart, allowing for the capture of multiple consecutive gait cycles in a controlled laboratory environment. We only used the straight-walking segments of the trials for our analysis. Compared to similar studies, a longer trial duration was intentionally chosen to improve the accuracy and reliability of both the mean and variability measures of gait parameters [61]. Although the extended duration could have induced fatigue, no participants reported feeling fatigued or requiring a break during the walking task. Gait data were recorded using a three-dimensional motion capture system (Vicon Nexus, version 2.3/2.8.2, Oxford Metrics, UK), consisting of 10 cameras and 61 reflective markers, sampled at 100 Hz.

### B. Gait parameter calculations

Gait parameters were computed using custom MATLAB scripts (version R2022a, The MathWorks Inc., Natick). In total, we selected 35 gait parameters, including the mean, variability, and asymmetry of common spatiotemporal parameters, along with measures of upper and lower limb coordination and margin of stability. The common spatiotemporal parameters include cadence (Cadence), double limb stance time (DLST), stride time (StrideT), stance time (StanceT), swing time (SwingT), step time (StepT), step length (StepL), step width (StepW), stride length (StrideL), and walking speed (WalkingSpeed). We computed the mean value of each common spatiotemporal parameter by averaging all individual steps recorded during the 10-minute walking trial. Asymmetry value (Asy) was evaluated for step length (StepL Asy), step width (StepW Asy), step time (StepT Asy), swing time (SwingT Asy), and stance time (StanceT Asy) [62]. Coordination between limbs was assessed using two established approaches: the Phase Coordination Index (PCI) [63] and Continuous Relative Phase (CRP) [13, 14]. PCI quantifies the consistency and accuracy of interlimb timing and was calculated for left–right step coordination (PCI LeftvsRight) and for coordination between short and long gait cycles (PCI ShortvsLong). CRP was used to assess the relative timing between limb segments and included coordination between upper limbs (CRP arm&arm), lower limbs (CRP Leg&Leg), and combined upper–lower limb interactions (CRP Rarm&Lleg, CRP Larm&Rleg, CRP Larm&Lleg, CRP Rarm&Rleg). The margin of stability was defined as the distance between the extrapolated center of mass and the boundaries of the base of support, defined by the toe markers (Metatarsal I and V), and was evaluated separately in the anteroposterior and medio-lateral directions [64–66]. The variability (Var) of each spatiotemporal parameter was quantified using the coefficient of variation, defined as (standard deviation / mean) × 100. The specific definition of each parameter is provided in Supplementary Methods 1. For common spatiotemporal parameters, foot clearance, and margin of stability, values from left and right gait cycles were averaged. Asymmetry and coordination parameters were computed for each gait cycle and then averaged over the whole trial after removing outliers, defined as values exceeding ±4 median absolute deviation (MAD) from the median.

To investigate the effects of DBS surgery on gait parameters, we conducted paired t-tests between the PRE and POST datasets for all 35 gait parameters, using a significance level of α=0.05. We also performed unpaired t-tests between the PRE dataset and age-matched health controls, using the same significance level. Gait parameters were analyzed individually to assess whether PD or DBS exerted parameter-specific effects. Therefore, no correction for multiple comparisons was applied in the statistical analysis.

### C. Classification framework

The classification framework comprised two primary objectives: first, to distinguish between PD patients and healthy controls; and second, to differentiate gait characteristics of patients before (PRE) and after (POST) DBS surgery. An identical workflow, comprising preprocessing, feature selection, classification, and evaluation, was applied to each classification objective. These steps are illustrated schematically in Figure 1.

**Fig. 1.**
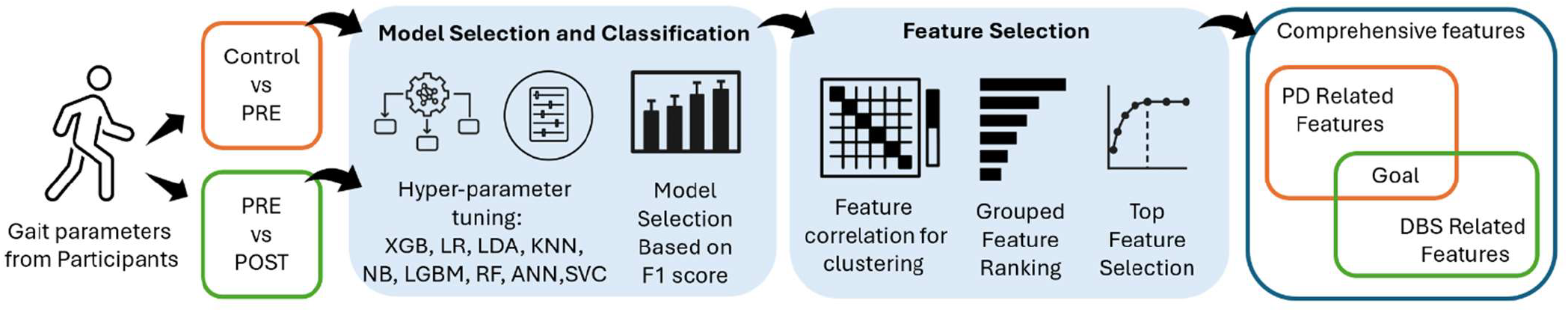
Overview of the gait-based classification and feature selection pipeline. Gait parameters were used to classify Control vs. PRE and PRE vs. POST conditions. Multiple machine learning models were tuned and evaluated based on the F1 score. Feature selection included correlation-based grouping and importance ranking. The final features were organized into PD-related and DBS-related subsets, with their overlap representing biomarkers that are both disease-relevant and treatment-responsive.

For the first classification objective, we used a cohort of 49 PwPD and 51 healthy controls. To ensure comparability between groups, we performed one-to-one nearest-neighbor matching based on age using the R package *MatchIt* [67]. Specifically, each patient was matched to a healthy control within a caliper of 0.5, effectively controlling for age-related confounding in our analysis.

After matching, we confirmed that there was no significant difference in age distribution between groups by performing a t-test with a significance level of α=0.05. This resulted in a final dataset of 44 PwPD and 44 age-matched healthy controls, which were combined and labeled accordingly. The data were then split into a training set (80%) and a test set (20%). The test set remained untouched throughout the model development and was only used in the final step to evaluate the optimized model with the selected features. Both the training and test sets were standardized separately using the mean and standard deviation of each feature. An extensive machine learning (ML) framework was implemented to systematically select, train, and evaluate several candidate models, as illustrated in Figure 1. Specifically, we included models previously demonstrated to be effective for similar classification tasks [49, 56, 57, 68, 69], such as XGBoost (XGB), logistic regression (LR), linear discriminant analysis (LDA), k-nearest neighbor (KNN), Naive Bayes (NB), support vector machine (SVC), random forest (RF), Light Gradient Boosting Machine (LGBM), and artificial neural networks (ANN).

For each ML model, we performed hyperparameter tuning using grid search optimization. Within this optimization framework, a 5-fold cross-validation approach was employed: the data were randomly divided into five subsets, stratified to maintain a consistent class balance (50%-50%) in each fold. During cross-validation, each hyperparameter combination was evaluated by training on four subsets and validating on the remaining subset. The hyperparameter set that yielded the highest average performance (measured F1-score) across the five validation folds was selected as optimal. Subsequently, the model with the highest classification performance among the 9 optimal models, determined primarily by the F1-score, was selected as the best-performing model.

Feature importances were extracted directly from this optimized, best-performing model, enabling the identification and ranking of the gait parameters that most strongly influenced classification outcomes.

In addition to distinguishing patients from healthy controls, we trained another set of models to differentiate between patients’ gait characteristics before and after DBS surgery. We applied the same modelling framework as before, with an added group-wise stratification to ensure that the PRE and the POST samples from the same patient were assigned exclusively to either the training or the test set, thereby preventing data leakage.

### D. Explainable AI–based gait feature selection

To identify the most critical gait parameters that differentiate PwPD from healthy controls, we employed an explainable AI approach using SHAP values to quantify feature contributions in a model-agnostic manner. In addition to the model’s built-in importance metrics, we employed SHAP values to gain deeper insights into feature contributions. SHAP is a consistent, model-agnostic interpretive framework that quantifies the marginal contribution of each feature to the model’s predictions, facilitating both global and individual-level explanations [70].

Before computing SHAP values, we evaluated Pearson’s correlation between individual features to minimize the impact of multicollinearity on model interpretability. Features with correlation coefficients greater than 0.9 were grouped together, whereas features without high correlation remained separate. SHAP values were subsequently calculated at the group level, determining each group’s overall importance in the classification process. These grouped SHAP rankings were used to guide the subsequent iterative feature selection.

We then adopted an iterative feature selection process, incrementally adding the top-ranked parameters and evaluating model performance at each step. At each iteration, we computed cross-validated F1-scores and identified the subset of gait parameters that yielded the highest classification accuracy. If multiple parameter subsets resulted in equally high accuracy, the smallest subset was selected as the optimal discriminative set. Finally, the entire training set was used to train the optimized model with the selected features, which was then applied to classify the test set. Classification performance was evaluated using the F1 score. The same methodology was applied to the second classification scenario, which focused on differentiating gait performance before and after DBS surgery.

## III. Results

### A. Participant Demographics

The original cohort comprised 51 healthy controls, who were each evaluated once, and 49 PwPD evaluated both PRE and POST surgery. After age-matching, 44 PwPD in the PRE condition were matched with 44 healthy controls, resulting in no statistically significant age difference between groups (P = 0.07). After selection, the average age value for PD is 60.50 (standard deviation: 10.90), while for healthy controls, it is 64.57 (standard deviation: 10.16). Notably, none of the participants exhibited freezing of gait during objective gait assessments; however, mild freezing of gait was self-reported by three participants prior to surgery and five participants following surgery, as indicated by their UPDRS scores. The details of the demographics include mean and standard deviation for age, weight, height, age at disease onset, disease duration, and disease severity according to the Hoehn and Yahr scale, as well as sex distribution are shown in Supplementary Table 1.

**TABLE 1.** The means (SD) of gait parameters for Pre-DBS, Post-DBS, and the control group are SHOWN, along with the corresponding p-values. Statistically significant differences are indicated in bold.

| Gait Parameters | Pre-DBS<br>(N=49) | Post-DBS<br>(N=49) | p-value<br>(pre-post) | Selected Pre-DBS<br>(N=44) | Selected Healthy<br>Control (N=44) | p-value<br>(pre-control) |
| --- | --- | --- | --- | --- | --- | --- |
| Cadence (steps/min) | 113.46 (7.74) | 115.8 (6.81) | **0.004** | 113.29 (7.66) | 116.64 (8.26) | 0.059 |
| DLST (s) | 0.13 (0.02) | 0.12 (0.02) | 0.255 | 0.13 (0.02) | 0.12 (0.02) | 0.358 |
| StrideT (s) | 1.06 (0.07) | 1.04 (0.06) | **0.004** | 1.06 (0.07) | 1.03 (0.08) | 0.065 |
| StanceT (s) | 0.66 (0.05) | 0.64 (0.05) | 0.014 | 0.66 (0.05) | 0.64 (0.05) | 0.107 |
| SwingT (s) | 0.41 (0.02) | 0.4 (0.02) | **0.005** | 0.41 (0.02) | 0.39 (0.02) | 0.035 |
| StepL (cm) | 64.49 (8.17) | 63.51 (8.28) | 0.214 | 64.93 (7.95) | 64.39 (5.48) | 0.702 |
| StepT (s) | 0.53 (0.04) | 0.52 (0.03) | **0.003** | 0.53 (0.04) | 0.52 (0.04) | 0.064 |
| StepW (cm) | 7.76 (2.95) | 8.19 (3.09) | 0.094 | 7.77 (2.97) | 8.16 (2.06) | 0.528 |
| StrideL (cm) | 127.82 (16.51) | 125.77 (16.65) | 0.196 | 128.71 (16.02) | 127.6 (11.14) | 0.697 |
| WalkingSpeed (cm/s) | 120.41 (19.07) | 121.29 (18.81) | 0.628 | 120.93 (18.47) | 123.88 (13.52) | 0.315 |
| StepL Asy (%) | 4.98 (3.17) | 5.47 (3.14) | 0.326 | 4.81 (3.04) | 3.01 (2.12) | **0.001** |
| StepW Asy (%) | 26.95 (11.47) | 23.01 (9.45) | **0.005** | 27.37 (11.68) | 20.94 (6.26) | **0.004** |
| StepT Asy (%) | 3.73 (2.63) | 4.4 (2.52) | 0.087 | 3.26 (1.41) | 2.53 (1.32) | **0.015** |
| StanceT Asy (%) | 2.72 (2.49) | 3.1 (2.21) | 0.14 | 2.36 (1.31) | 1.91 (1.0) | 0.099 |
| SwingT Asy (%) | 3.89 (3.82) | 4.5 (3.4) | 0.128 | 3.36 (2.31) | 2.75 (1.61) | 0.174 |
| PCI LeftvsRight (%) | 181.21 (3.9) | 180.67 (4.43) | 0.385 | 180.5 (2.88) | 180.35 (2.29) | 0.798 |
| PCI ShortvsLong (%) | 181.02 (3.98) | 180.47 (4.43) | 0.38 | 180.3 (2.91) | 180.17 (2.28) | 0.817 |
| CRP Leg&Leg (unitless) | 0.87 (0.21) | 0.87 (0.23) | 0.894 | 0.87 (0.25) | 0.76 (0.17) | **0.013** |
| CRP arm&arm (unitless) | 0.88 (0.21) | 0.88 (0.23) | 1 | 0.88 (0.24) | 0.77 (0.17) | **0.008** |
| CRP Rarm&Lleg (unitless) | 0.84 (0.26) | 0.73 (0.1) | **0.003** | 0.84 (0.23) | 0.69 (0.08) | **<0.001** |
| CRP Larm&Rleg (unitless) | 0.87 (0.25) | 0.75 (0.1) | **0.003** | 0.87 (0.23) | 0.72 (0.08) | **<0.001** |
| CRP Rarm&Rleg (unitless) | 2.63 (0.24) | 2.55 (0.08) | 0.018 | 2.61 (0.23) | 2.53 (0.08) | **0.037** |
| CRP Larm&Lleg (unitless) | 2.61 (0.24) | 2.53 (0.08) | 0.018 | 2.59 (0.23) | 2.51 (0.08) | **0.037** |
| MOS AP (mm) | -29.2 (27.01) | -22.52 (14.25) | 0.142 | -31.34 (26.47) | -17.84 (16.83) | **0.007** |
| MOS ML (mm) | 69.61 (25.5) | 68.48 (22.02) | 0.810 | 71.11 (24.45) | 76.8 (19.05) | 0.196 |
| Cadence Var (% CV) | 1.64 (0.68) | 1.82 (0.86) | 0.179 | 1.57 (0.57) | 1.54 (0.86) | 0.871 |
| DLST Var (% CV) | 8.29 (2.39) | 9.26 (3.34) | 0.067 | 8.08 (2.14) | 6.75 (2.59) | **0.018** |
| StrideT Var (% CV) | 1.96 (0.61) | 2.15 (0.82) | 0.141 | 1.89 (0.52) | 1.82 (0.86) | 0.641 |
| StanceT Var (% CV) | 2.7 (0.74) | 3.01 (1.02) | 0.042 | 2.6 (0.62) | 2.42 (1.06) | 0.367 |
| SwingT Var (% CV) | 2.54 (0.68) | 2.78 (1.02) | 0.096 | 2.49 (0.66) | 2.03 (0.78) | **0.006** |
| StepL Var (% CV) | 3.52 (1.05) | 3.76 (1.31) | 0.177 | 3.4 (0.97) | 2.59 (0.64) | **<0.001** |
| StepT Var (% CV) | 2.44 (0.59) | 2.64 (0.91) | 0.134 | 2.37 (0.51) | 2.16 (0.8) | 0.167 |
| StepW Var (% CV) | 32.69 (15.3) | 28.3 (12.97) | **0.017** | 33.24 (15.56) | 23.54 (7.74) | **0.002** |
| StrideL Var (% CV) | 3.11 (1.07) | 3.32 (1.27) | 0.222 | 2.96 (0.96) | 2.32 (0.73) | **0.002** |
| WalkingSpeed Var (% CV) | 4.0 (2.67) | 3.91 (1.49) | 0.816 | 3.87 (2.61) | 3.15 (1.46) | 0.128 |

### B. Comparative analysis of gait parameters across PD and DBS states

The average and standard deviation values of all 35 gait parameters for each group, as well as the P values from the t-tests, are shown in Table 1. By comparing PwPD and healthy conditions, 16 out of 35 gait characteristics were significantly affected by DBS treatment. These include SwingT, StepL Asy, StepW Asy, StepT Asy, CRP Leg&Leg, CRP arm&arm, CRP Rarm&Lleg, CRP Larm&Rleg, CRP Rarm&Rleg, CRP Larm&Lleg, MOS AP, DLST Var, SwingT Var, StepL Var, StepW Var, and StrideL Var. On the other hand, for the comparison between PRE and POST conditions, 11 out of 35 gait characteristics were significantly impaired in PD. These include Cadence, StrideT, SwingT, StepT, StepW Asy, CRP Rarm&Lleg, CRP Larm&Rleg, CRP Rarm&Rleg, CRP Larm&Lleg, StanceT Var, and StepW Var.

### C. Classification performance

All 35 gait parameters were used as input features. Figure 2 presents the cross-validated F1-scores on the validation set for nine models in both classification scenarios, with XGBoost yielding the highest performance in each.

**Fig. 2.**
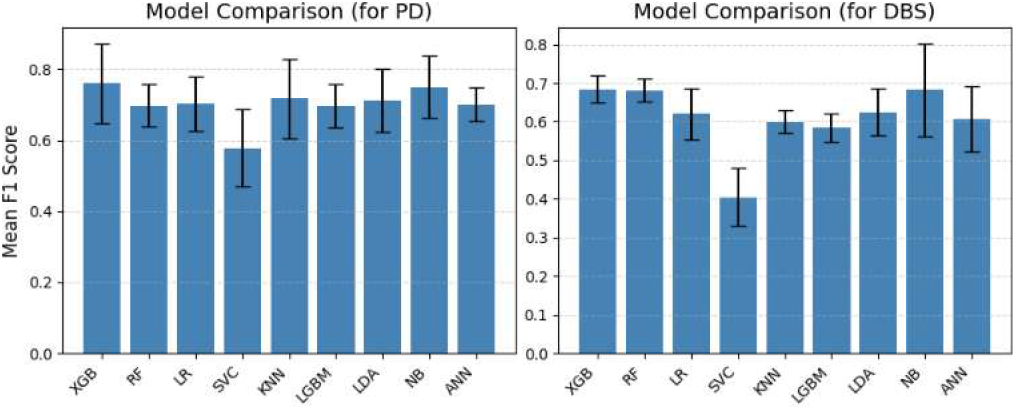
The performance of the models on the classification tasks: 1. distinguishing PwPD from healthy controls (left); and 2. distinguishing PRE and POST surgery gait in PwPD (right). F1 scores are used to evaluate classification performance. Mean values and std deviations after 5-fold cross-validation were shown in the bar plots.

We subsequently selected the XGBoost model as the primary classifier for our parameter selection approach. For the classification between healthy controls and PwPD before DBS treatment, the F1 score for the training set was 0.92 ± 0.03, while the F1 score for the validation set was 0.76 ± 0.11 (Table 2). For the classification between PRE and POST, the F1 score for the training set was 0.85 ± 0.06, while the F1 score for the validation set was 0.69 ± 0.06.

**TABLE 2.**
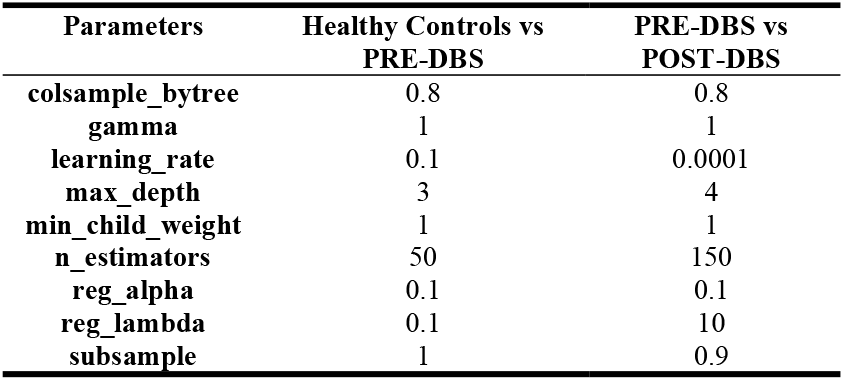
The best-performing hyperparameters for the XGBoost model.

| Parameters | Healthy Controls vs PRE-DBS | PRE-DBS vs POST-DBS |
| --- | --- | --- |
| colsample_bytree | 0.8 | 0.8 |
| gamma | 1 | 1 |
| learning_rate | 0.1 | 0.0001 |
| max_depth | 3 | 4 |
| min_child_weight | 1 | 1 |
| n_estimators | 50 | 150 |
| reg_alpha | 0.1 | 0.1 |
| reg_lambda | 0.1 | 10 |
| subsample | 1 | 0.9 |

Several of the selected gait features exhibited strong pairwise correlations (Figure 3). As expected, several features showed strong pairwise correlations, with correlation coefficients approaching 1.0. Notably, features representing different statistical descriptors of the same gait metric, such as mean and variability, or mean and asymmetry, tended to cluster together with high correlations. Similarly, parameters describing left– right and cross-limb coordination (e.g., CRP groups) also demonstrated strong intercorrelations. To address this redundancy and improve interpretability, we grouped together features with correlation coefficients greater than 0.9, resulting in 20 feature clusters. These clusters were subsequently used to compute grouped SHAP values, with detailed information provided in Supplementary Table 2.

**Fig. 3.**
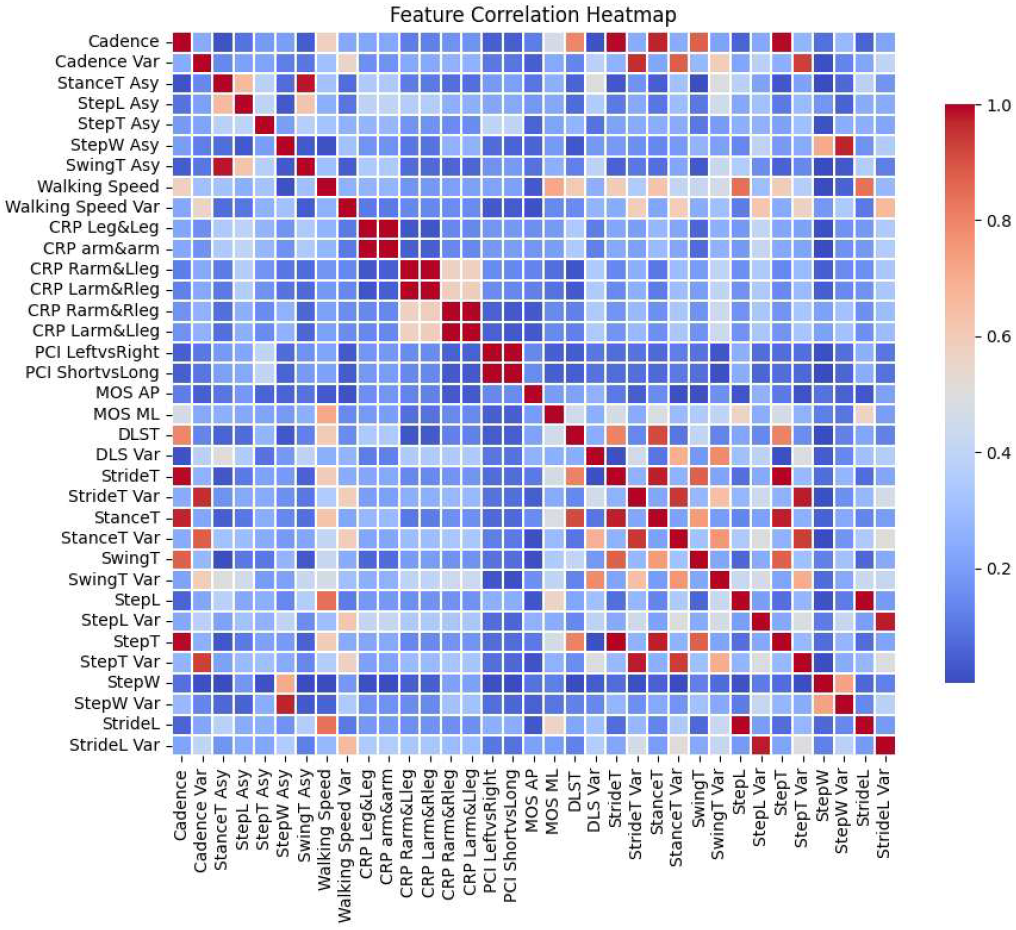
The heatmap shows the correlations across the 35 features. The values represent the absolute value of the Pearson correlation coefficient. Dark blue indicates no correlation (correlation = 0), while dark red indicates perfect correlation (correlation = 1), meaning the two features vary identically.

The explainable AI analysis based on grouped SHAP values revealed the grouped feature importance rankings, quantified as mean absolute SHAP values, for the PD versus healthy control classification (Figure 4 left) and the pre-versus post-DBS classification (Figure 4 right). In the grouped SHAP plots, dark blue bars represent the most representative feature within each correlation cluster, while the adjacent light blue bars indicate other features within the same cluster (e.g., StepW Asy and StepW Var).

**Fig. 4.**
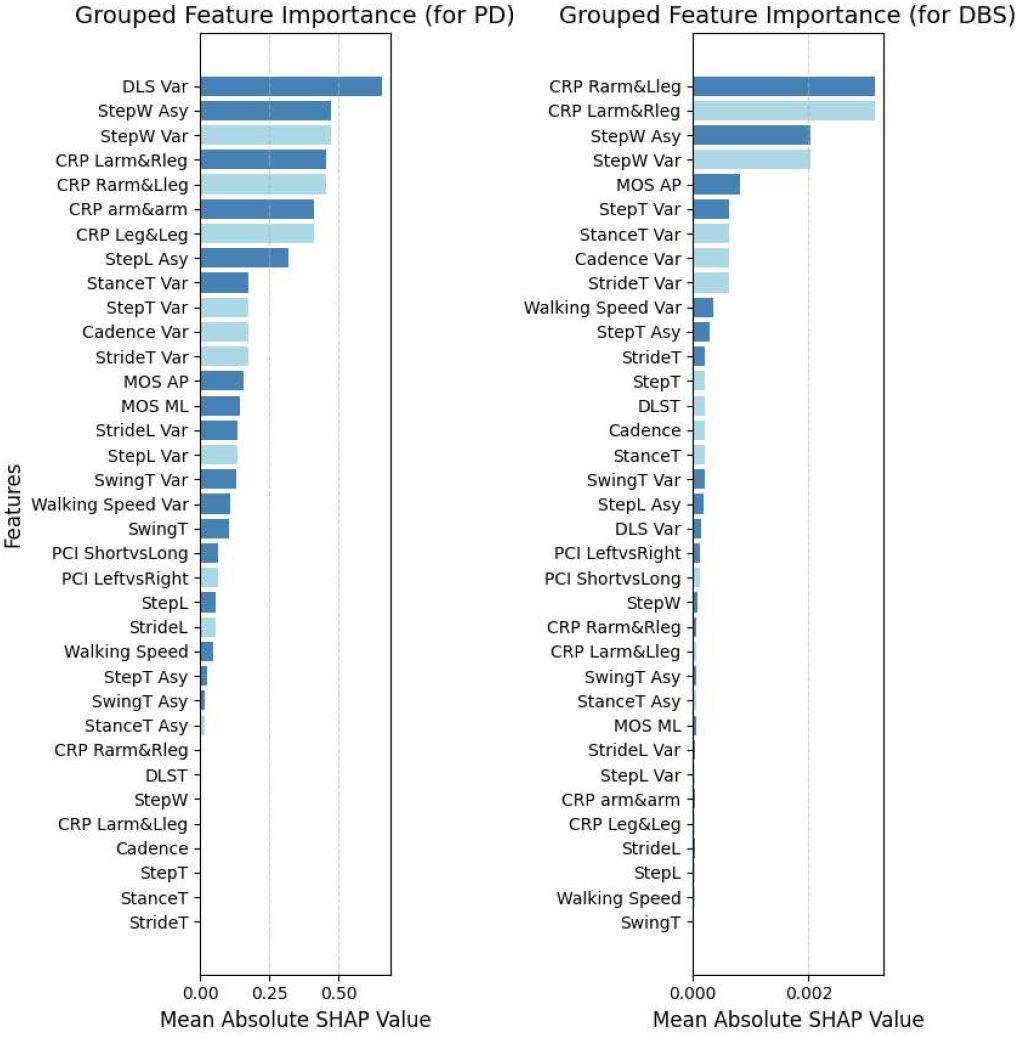
The feature importance ranking of all 35 gait features for PD (left) and DBS (right), which represents the importance of the feature to understand PD or DBS-related effects on patients. Feature importance is represented by the mean absolute SHAP values; higher SHAP values indicate greater influence on the model’s predictions. Features within the same correlation group share the same SHAP value, with the most representative feature shown in dark blue and the remaining group members in light blue. Features are ranked from top to bottom in order of decreasing importance. The top-ranking features are the most important for understanding PD or DBS-related effects on patients.

In the PD classification task, gait variability and asymmetry emerged as the most discriminative domains, led by double limb stance time variability (DLS Var), step width asymmetry (StepW Asy), and step width variability (StepW Var), followed by bilateral coordination measures such as CRP Larm&Rleg, CRP Rarm&Lleg, CRP arm&arm, and CRP Leg&Leg.

In contrast, the DBS classification prioritized cross-limb coordination (CRP Rarm&Lleg, CRP Larm&Rleg), step width asymmetry, step width variability, and anteroposterior margin of stability (MOS AP). Notably, five features, StepW Asy, StepW Var, CRP Larm&Rleg, CRP Rarm&Lleg, and MOS AP, were consistently ranked among the most important in both tasks.

### D. Parameter selection based on model performance

We assessed the F1-score while incrementally including top-ranked parameters to identify the gait parameters most influenced by PD and DBS treatment. Figure 5 depicts the relationship between classification performance (F1 score) and the number of selected parameters. For the PD-related classification (PwPD vs. healthy controls), the classifier achieved its highest performance (F1 score: 0.81 ± 0.07) when utilizing the top 13 parameters: DLS Var, StepW Asy, StepW Var, CRP Larm&Rleg, CRP Rarm&Lleg, CRP arm&arm, CRP leg&leg, StepL Asy, StanceT Var, StepT Var, Cadence Var, StrideT Var, and MOS AP. The performance of the optimized model with selected features on classifying the test set is similar (F1 score = 0.75).

**Fig. 5.**
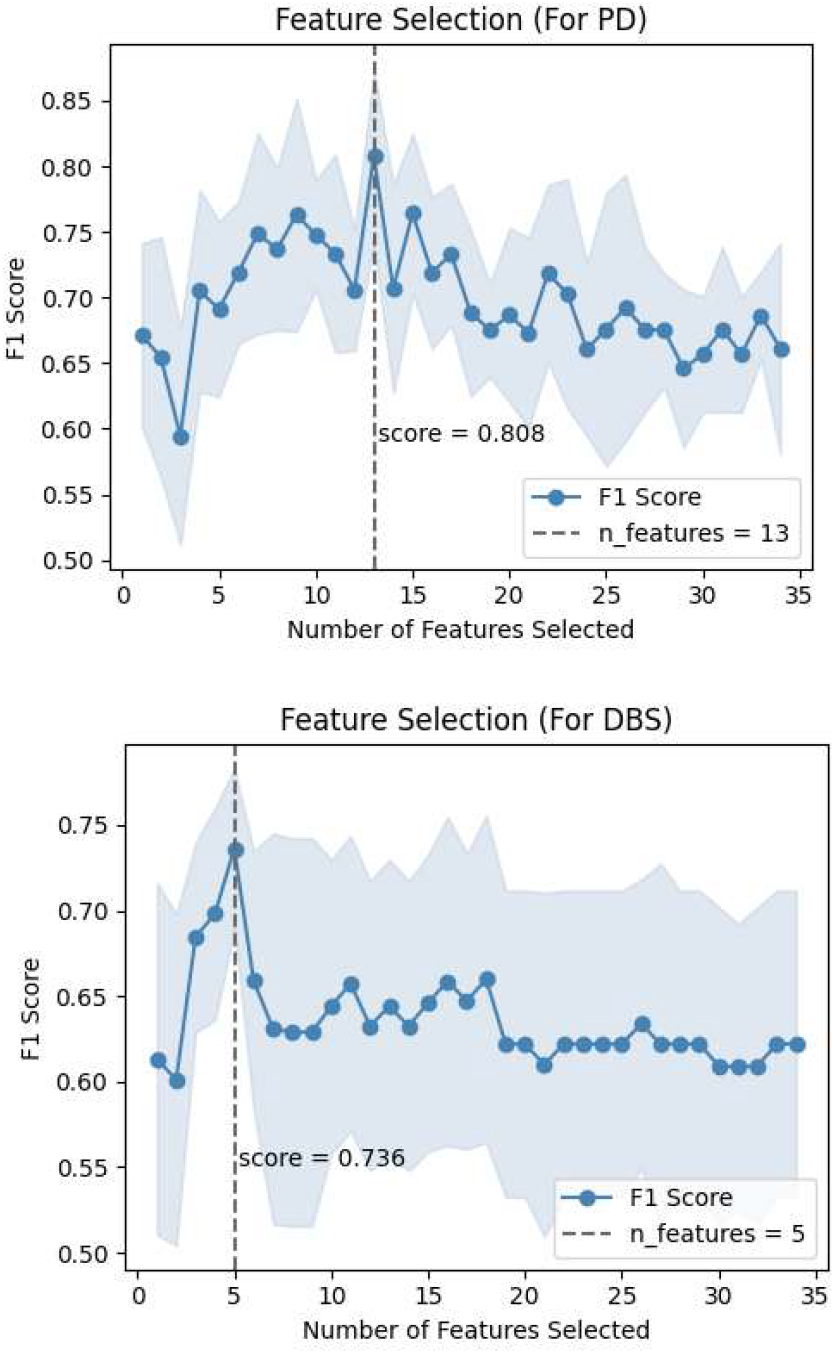
The model’s performance when selecting a different number of the most related gait parameters as the input for PD (up) and DBS (down). The x-axis represents the number of top-ranked features used as input, while the y-axis shows the average F1 score across 5-fold cross-validation (mean ± standard deviation). The dashed line marks the iteration where the model achieved its peak performance, indicating the optimal number of gait features for characterizing the effects of PD and DBS, respectively.

When differentiating gait before and after DBS surgery, the classifier reached optimal performance (F1 score: 0.74 ± 0.05) using the top 5 parameters: CRP Rarm&Lleg, CRP Larm&Rleg, StepW Asy, StepW Var, and MOS AP. The performance of the optimized model with selected features on the test set using these selected features was similar (F1 score = 0.72).

## IV. Discussion

This study integrated statistical analysis, machine learning, and explainable AI to identify gait parameters that distinguish PD from healthy controls and pre-from post-DBS conditions. Statistical tests revealed group differences, while model-based ranking and selection highlighted parameters with optimized predictive value. Grouped SHAP analysis provided biomechanically interpretable explanations, resulting in feature sets that capture both distinct and overlapping gait characteristics associated with disease and stimulation effects. By systematically identifying gait parameters that both characterize PD-related deficits and respond to DBS, this study addresses a critical gap in the field and provides a framework for linking biomechanical markers with therapeutic outcomes.

### A. The influence of PD and DBS on gait parameters

Statistical comparisons revealed that medio-lateral stability measures, particularly step width variability and step width asymmetry, were significantly higher in PD than in healthy controls. These abnormalities are consistent with impaired lateral balance control caused by deficits in proprioceptive feedback, as well as asymmetrical motor output that reflects the unilateral onset and progression of PD [71, 72]. Temporal variability measures, including double-support time and stance phase variability, were also elevated, indicating reduced consistency in gait cycle timing and diminished rhythmic motor control by basal ganglia cortical circuits [73–75].

Following DBS, reductions in step width variability and double-support time variability indicated partial restoration of stability control and rhythmicity [44]. Improvements in these parameters are consistent with evidence that subthalamic stimulation can modulate subcortical–cortical and brainstem pathways involved in postural control [39, 48, 76]. However, step width asymmetry persisted after DBS, suggesting that symmetry-related control mechanisms, which are thought to be less responsive to dopaminergic or stimulation-based modulation, remain impaired. This aligns with reports that axial symptoms are resistant to current pharmacological and surgical interventions in PD [77]. Beyond their established role as fall-risk indicators, our results also suggest that step width metrics are sensitive to DBS-related improvements, highlighting their potential dual role as markers of both disease burden and therapeutic efficacy.

### B. Explainable AI framework and model choice

Nine classifiers from different methodological families were evaluated to avoid bias toward a single modeling approach and to compare their suitability for structured gait data. XGBoost achieved the highest cross-validated F1-scores on the validation set for both classification tasks. This aligns with findings from other structured biomedical datasets, where gradient boosting often outperforms linear models and kernel-based methods [78]. Its advantage here is likely due to its ability to capture nonlinear relationships among heterogeneous gait parameters, built-in regularization through L1/L2 penalties, and robustness to skewed or interdependent variables [79].

Although hyperparameters were tuned with a focus on generalization, the gap between training and test F1-scores indicates residual overfitting. The shallow tree depth and small learning rate constrained model complexity, while subsampling observations and features reduced correlation between trees. However, the limited dataset size likely prevented further reduction of variance without loss of predictive power [80]. The stability of validation scores across folds suggests that the observed overfitting is a data limitation rather than a tuning issue.

For interpretability, SHAP was chosen over alternatives such as permutation importance due to its theoretical guarantees of consistency and local accuracy, allowing importance scores to be interpreted at both the population and individual levels [81]. To address multicollinearity, features with Pearson correlation coefficients above 0.9 were grouped before SHAP computation, ensuring that each score represented a distinct biomechanical construct rather than redundant metrics. While this approach reduces resolution for highly correlated features, it enhances the robustness and clinical interpretability of the importance rankings. The main limitation of SHAP, its computational demands and dependence on training data distribution, was mitigated through feature grouping and repeated cross-validation, improving stability of the explanations.

### C. Interpretation of selected gait parameters for PD and DBS

The feature selection process yielded two distinct sets: for PD classification, parameters were primarily related to medio-lateral stability deficits, bilateral coordination, and temporal control. While some prior studies identified variability as the most discriminative marker for PD [52, 56]. others highlighted asymmetry and temporal parameters as equally important [54]. For DBS classification, the retained features emphasized medio-lateral and anteroposterior stability and cross-limb coordination. Notably, several coordination features derived from CRP were consistently retained. Although less frequently emphasized in earlier gait studies, CRP has been proposed as a sensitive marker of impaired interlimb coupling in PD, which aligns with its prominence in our feature selection [82]. Although group-level comparisons showed partial improvements in temporal variability measures after DBS, these features were not retained by the classification models, suggesting that their responsiveness is inconsistent across individuals and less reliable as markers of stimulation effects.

The F1 scores for the final feature subsets were lower on the validation set than in training, reflecting a degree of overfitting. Across repeated cross-validation folds, performance showed moderate variability, with PRE vs. POST classification yielding relatively consistent results compared to the larger fluctuations observed in HC vs. PRE-DBS. This pattern suggests that while the selected features contributed meaningfully to model discrimination, their stability across samples is only partial and limited by the dataset size. Nevertheless, the convergence of statistical group differences, model performance, and biomechanical plausibility supports the credibility of the identified features.

### D. Overlap between the most affected gait parameters

Five gait parameters, step width asymmetry, step width variability, coordination between the left arm and right leg, coordination between the right arm and left leg, and the anteroposterior margin of stability, emerged as critical overlapping indicators of PD-related impairments and DBS responsiveness. All five shifted toward healthy control values after DBS, indicating improvements in medio-lateral stability, interlimb coordination, and forward–backward balance.

From a neurophysiological perspective, bilateral coordination deficits in PD arise from basal ganglia dysfunction, which disrupts motor planning, sequencing, and the timing of arm-leg movements [83]. Optimized bilateral DBS stimulation may partially restore these functions by modulating basal ganglia-cortical and brainstem pathways, thereby improving rhythmicity and promoting more symmetrical movement patterns [84]. Similarly, elevated step width variability and asymmetry may reflect impaired internal rhythm and timing control, resulting in inconsistent foot placement and unstable gait [85]. This interpretation is consistent with models of PD gait showing that deficits in pace, rhythm, variability, and asymmetry are key hallmarks of impaired locomotor control.

Interestingly, in our study, the mean values of basic spatiotemporal parameters did not emerge as important in either classification task. Although previous studies have reported mean-based parameters, such as step length or stance duration, as discriminative [39–44]. In our dataset, these measures were not retained after feature selection, possibly because averaging could reduce subtle but consistent gait abnormalities over a prolonged 10-minute continuous walking protocol. In contrast, variability- and asymmetry-based measures, which accumulate diagnostic value over longer recordings, provide a more robust representation of intrinsic PD pathology and DBS-induced improvements.

As a group, these five features offer a compact, physiologically grounded set of markers that can help clinicians objectively assess gait impairments, quantify the impact of DBS, and monitor disease progression in PD. Their combined sensitivity to both pathological deficits and stimulation-induced improvements means they could be incorporated into standardized gait assessment protocols, enabling more precise treatment evaluation, personalized rehabilitation planning, and early detection of changes that may require adjustments to DBS parameters or additional therapeutic interventions.

### E. Limitations

This study has several limitations. First, increasing the cohort size could lead to better classification accuracy and more reliable feature importance ranking. A limited sample size also increases the risk of overfitting, where the model captures dataset-specific patterns that may not generalize well to broader populations. Secondly, only optimized bilateral DBS settings were examined, leaving open the question of how different stimulation parameters or real-world walking contexts influence the identified markers.

## V. Conclusion

This study provides the first comprehensive classification of gait parameters that both characterize PD-related motor impairments and are sensitive to DBS-induced improvements, within an interpretable machine learning framework. By combining statistical comparisons with model-based feature selection and SHAP-based interpretation, we identified a compact set of gait parameters that not only distinguished PD from healthy gait but also reflected stimulation-induced improvements. The consistent emergence of step width metrics, interlimb coordination measures, and the anteroposterior margin of stability underscores their central role in both disease-related impairment and therapeutic response.

These findings suggest that a small, physiologically grounded set of gait parameters can serve as objective biomarkers to evaluate DBS effects and track disease progression in PD. With validation in larger cohorts, these markers could be integrated into clinical assessment and monitoring pipelines, supporting more precise treatment decisions and individualized rehabilitation strategies.

## Data Availability

All data produced in the present study are available upon reasonable request to the authors.

## Acknowledgment

The authors extend their gratitude to all project members involved in the original data collection, including Dr. Navrag Singh, Prof. Dr. William Taylor, Dr. Christian Baumann, and their respective team members.

