## Supplementary material for "Using Explainable AI to Identify Disease-Relevant and Deep Brain Stimulation Treatment-Sensitive Gait Features in Parkinson’s Disease": Supp. Methods 1

**Supp. Method 1: Parameter Identification**

**Cadence** is the number of steps a person takes per minute while doing overground walking.

**Double limb support time** is the period of time when both feet are in contact with the ground.

**Stride Time** is the time elapsed between the heel contact of one foot and the heel contact of the ipsilateral foot when it touches the ground again.

**Stance time** is the time elapsed between heel contact of one foot and toe-off of the ipsilateral foot when it leaves the ground.

**Swing time** is the time elapsed between the toe-off of one foot and the heel contact of the ipsilateral foot when it touches the ground again.

**Step length** is the anterior-posterior distance between the heel contact of one foot and the heel contact of the contralateral foot.

**Step time** is the time elapsed between heel contact of one foot and heel contact of the contralateral foot.

**Step width** is the medial-lateral distance between the place of one foot and the center lines of the two feet.

**Stride length** is the anterior-posterior distance between the heel contact of one foot and the heel contact of the ipsilateral foot.

**Walking speed** is evaluated as the ratio of the distance covered by the sacrum marker between two consecutive gait cycles to the ambulation time.

**Maximum Heel Clearance** is the greatest vertical distance between the marker placed on the calcaneus of the foot and the walking surface during the swing phase of human gait.

**Minimum Toe Clearance** is the smallest vertical distance between the marker placed on the third metatarsal and the walking surface during the swing phase of human gait.

**Margin of Stability** is defined as the distance between the extrapolated center of mass and the boundaries of the base of support, defined by the toe markers (Metatarsal I and V)

**Asymmetry** is the relative difference between the left and right gait values, normalized to the larger value.

**PCI** (expressed in percentage) is the sum of the coefficient of variation of the phase and the mean absolute difference between the phase and 180°. The relative timing of contralateral heel contacts is defined as the phase.

**PCI_LeftvsRight** is the PCI between the left and right stepping phases

**PCI_ShortvsLong** is the PCI between the previously labelled short leg (the limb with the smaller swing phase in that stride) and long leg.

**CRP** is derived by applying a Hilbert transform to the normalized angle trajectories from two selected limbs to obtain their instantaneous phases and then calculating the wrapped phase difference across the gait cycle, providing a continuous measure of interlimb coordination.

**CRP_Leg&Leg** is the CRP between two lower limbs.

**CRP_arm&arm** is the CRP between the two upper limbs.

**CRP_Rarm&Lleg** is the CRP between the right upper limb and the left lower limb.

**CRP_Larm&Rleg** is the CRP between the left upper limb and the right lower limb.

**CRP_Rarm&Rleg** is the CRP between the right upper limb and the right lower limb.

**CRP_Larm&Lleg** is the CRP between the left upper limb and the left lower limb.
