## Supplementary material for "Using Explainable AI to Identify Disease-Relevant and Deep Brain Stimulation Treatment-Sensitive Gait Features in Parkinson’s Disease": Supp. Table 1

**Supp. Table 1: Baseline and Longitudinal Demographic Characteristics of the original cohort**

| **Characteristics** | **Healthy Controls (N = 51)** | **PwPD Pre-DBS (N = 49)** | **PwPD Post-DBS (N = 49)** |
|---|---|---|---|
| Age (years) | 66.6 (10.7) | 59.7 (10.4) | - |
| Male/Female (No.) | 22/29 | 41/8 | - |
| Weight (kg) | 68.1 (12.2) | 76.8 (12.6) | 78.8 (11.9) |
| Height (cm) | 168.9 (8.8) | 176.1 (6.8) | 175.7 (7.0) |
| Age at disease onset (years) | - | 50.8 (9.7) | - |
| Disease duration (years) | - | 8.9 (4.5) | - |
| H&Y stage | - | 2 (1–3) | - |
| MDS-UPDRS I | - | 9.76 (5.1) | 7.0 (3.3) |
| MDS-UPDRS II | - | 12.76 (4.3) | 8.0 (6.4) |
| MDS-UPDRS III | - | 18.9 (6.9) | 17.0 (7.9) |
| MDS-UPDRS IV | - | 7.62 (4.1) | 0.4 (2.6) |
| LEDD [mg] | - | 1060 (473.5) | 347.8 (295.9) |

*Values are expressed as numbers of participants or mean (standard deviation). Baseline demographics between the two groups (PwPD Pre-DBS and Healthy Controls) were significantly different (P<0.01, two-sample t-tests and chi-squared tests). Abbreviations: PwPD, patients with Parkinson's disease; DBS, deep brain stimulation; H&Y, Hoehn and Yahr.*
