## Supplementary material for "Using Explainable AI to Identify Disease-Relevant and Deep Brain Stimulation Treatment-Sensitive Gait Features in Parkinson’s Disease": Supp. Table 2

**Supp. Table 1: Parameter Clusters**

| Groups | Parameters in the group |
|---|---|
| 1 | 'SwingT Asy', 'StanceT Asy' |
| 2 | 'CRP arm&arm', 'CRP Leg&Leg' |
| 3 | 'CRP Larm&Rleg', 'CRP Rarm&Lleg' |
| 4 | 'CRP Larm&Lleg', 'CRP Rarm&Rleg' |
| 5 | 'PCI ShortvsLong', 'PCI LeftvsRight' |
| 6 | 'StrideT', 'Cadence', 'StepT', 'StanceT', 'DLST' |
| 7 | 'StrideT Var', 'StanceT Var', 'StepT Var', 'Cadence Var' |
| 8 | 'StepL', 'StrideL', 'Walking Speed' |
| 9 | 'StepW Var', 'StepW Asy' |
| 10 | 'StrideL Var', 'StepL Var' |
| 11 | 'StepW' |
| 12 | 'MOS ML' |
| 13 | 'MOS AP' |
| 14 | 'Walking Speed Var' |
| 15 | 'StepT Asy' |
| 16 | 'DLS Var' |
| 17 | 'SwingT Var' |
| 18 | 'StepL Asy' |
| 19 | 'SwingT' |
